# Cannabidiol for treatment of Irritability and Aggressive Behavior in Children and Adolescents with ASD: Background and Methods of the CAnnabidiol Study in Children with Autism Spectrum DisordEr (CASCADE) Study

**DOI:** 10.1101/2024.08.12.24311894

**Authors:** Elise M. Sannar, Joan R. Winter, Ronda K. Franke, Emily Werner, Rebecca Rochowiak, Patrick W. Romani, Owen S. Miller, Jacquelyn L. Bainbridge, Obehi Enabulele, Talia Thompson, Crystal Natvig, Susan K. Mikulich-Gilbertson, Nicole R. Tartaglia

## Abstract

**Introduction:** Autism spectrum disorder (ASD) is a neurodevelopmental disorder commonly associated with behavioral challenges. There are few evidence based pharmacological interventions available for the treatment of behavioral symptoms associated with ASD. Cannabidiol (CBD), the non-psychoactive component of cannabis, has potential neuroprotective, antiepileptic, anxiolytic, and antipsychotic effects and may be useful in treating the behavioral symptoms of ASD.

**Methods:** We describe the research methods of a 27-week double-blind placebo-controlled cross-over trial of cannabidiol for the treatment of irritability and aggression associated with ASD, utilizing the irritability subscale of the Aberrant Behavior Checklist-2nd edition (ABC-2) as the primary outcome measure. Adverse effects and safety monitoring protocols are included. Several secondary and exploratory outcomes measures also include anxiety, communication, repetitive behaviors, attention, hyperactivity, autism family experience, and telehealth functional behavior assessment.

**Conclusion:** There is a significant need for clinical research exploring alternative medications for the treatment of behavioral symptoms of ASD. Cannabidiol (CBD) is being studied for the management of irritability, aggression, and other problem behaviors associated with ASD.

## INTRODUCTION

Autism spectrum disorder (ASD) is a neurodevelopmental condition characterized by deficits in social communication and restricted, repetitive patterns of behavior, interests, or activities. As described by the Diagnostic and Statistical Manual for Mental Disorders (DSM-5), the social communication deficits of ASD include impaired use of nonverbal communication, decreased reciprocity in social interactions and conversation, and lack of ability to read social cues or the body language of others. Additional criteria include inflexible adherence to routines, intense or restricted interests, sensory sensitivities, and stereotyped behaviors.[1] Beyond the core symptoms of ASD, many individuals have behavioral challenges including irritability, aggression, self-injury, anxiety, and sleep dysregulation.[2]

The prevalence of ASD has increased over the past few decades with current estimates from the United States Centers for Disease Control of 1 case in every 36 children.[3] However, available treatments for ASD have not significantly changed, and there is no medication treatment for the core symptoms of ASD. The only medications approved by the US Food and Drug Administration (FDA) for irritability and aggression associated with ASD are aripiprazole and risperidone.[4,5] While shown to be effective in Phase III clinical trials, these medications come with a high risk of side effects, including risk of diabetes, hypertension, weight gain, elevated triglycerides and movement disorders such as tardive dyskinesia.[6] Further, these medications are only effective for a segment of the population, and many patients continue to have sub-optimal benefit, leading to high risk of polypharmacy.[7,8] Incomplete treatment response and family preferences for complementary and alternative approaches have led to great interest in cannabinoid products as a potential treatment for symptoms of ASD.

Cannabidiol (CBD) is a non-intoxicating molecule found in strains of the cannabis or hemp plants which demonstrates neuroprotective, antiepileptic, anxiolytic, and antipsychotic properties by interacting with the body’s endogenous cannabinoid receptors and pathways.[9] There are multiple proposed ways that CBD may exert its effects on the endocannabinoid system, including modulation of neurotransmitter levels, neurotransmitter binding, cell signaling, and neurogenesis, and/or by altering circulating levels of endocannabinoids.[9] CBD has low affinity for the two known types of cannabinoid receptors (CB1R and CB2R). CB1R is primarily found in the central and peripheral nervous system, where it aids in migration and differentiation of neural progenitor cells in early development.[9,10] It is also involved in pathways regulating memory, cognition, and social function.[9,10] CB2R is predominately found in peripheral organs and tissues, such as the spleen and thymus, where it serves an immunomodulatory function. It is also found in neural networks during early development, where it contributes to CB1R effects in regulation of the cell cycle.[10] In addition to action at the CB1 and CB2 receptors, CBD acts on 5-HT1A serotonin [11] and TRPV1 pathways [12], inhibits adenosine reuptake [13,14], and regulates activity of PPAR-gamma homeostatic and metabolic regulatory pathways.[15]

Studies have demonstrated a potential role of the endocannabinoid system in the pathogenesis and treatment of ASD.[16–18] Human and mouse studies indicate alterations in both endocannabinoid ligands and receptors in people with autism as compared to controls. In mouse models with social impairments similar to ASD, there are improvements in social function with blockade of CB1R and fatty acid amide hydrolase (FAAH), the enzyme that hydrolyzes the primary endocannabinoid anandamide and other related endocannabinoid substances.[19] Post mortem brain analyses of individuals with ASD have found down-regulated CB1R expression.[20] Meanwhile, peripheral CB2R has been found to be upregulated in peripheral blood mononuclear cells of children with ASD.[21] Human studies have also found that children with ASD have lower levels of plasma anandamide and other endocannabinoids as compared to controls.[22,23]

There are multiple anecdotal reports, retrospective and open label trials showing improvements in the behavioral features of ASD with CBD treatment. The majority of these investigations include the use of CBD rich cannabis, rather than pure CBD, and thus cannot be generalized to the effects of CBD alone.[24–30]. The potential concern with these investigations is that CBD rich cannabis can also contain small amounts of THC (the psychoactive ingredient of the marijuana plant) which can lead to poor tolerability, adverse psychoactive effects, and are often not desired by parents and caregivers of children with ASD. Further, potential long term negative effects of THC on brain development are a common concern.[31,32]

A more recent placebo-controlled trial used pharmaceutically manufactured CBD topical gel to treat patients with Fragile X syndrome, the most common inherited genetic cause of ASD. [33–35] The trial showed decreased irritability and improved social interactions in the subset of participants with full methylation of the Fragile X gene, and the intervention was well tolerated. The results of these trials indicate a need for further research, including a placebo-controlled trial of pure CBD in patients with ASD.

The pharmaceutical formulation of plant-derived, highly purified cannabidiol (CBD) is FDA approved as Epidiolex ® in the United States for seizures associated with three rare epilepsy syndromes (Lennox Gastaut syndrome, Dravet syndrome, and Tuberous Sclerosis Complex) in patients ≥ 1 year.[36] Multiple double-blind, placebo-controlled studies demonstrated the general safety profile of Epidiolex ® in pediatric and adult populations with developmental and epileptic encephalopathies populations with neurodevelopmental disorders, and efficacy in decreasing seizure frequency.[37–40]

In this report we describe methods of a double-blind, placebo-controlled study to evaluate the efficacy and safety of cannabidiol (CBD) for the treatment of irritability and aggression in youth with ASD.

## METHODS

### Overall Study Design

This is a single-site, double-blind, randomized, placebo-controlled study with modified cross-over design to evaluate the efficacy and safety of oral cannabidiol (CBD) for the treatment of irritability and aggression in youth aged 5-17 years with autism spectrum disorder.

There are three treatment arms (A, B, and C, respectively). Arm A receives CBD for the first twelve weeks (Period 1), followed by a three-week washout period, and then 12 weeks of placebo (Period 2). Arm B receives placebo for the first twelve weeks followed by a three-week washout period and then 12 weeks of active study drug. Arm C receives CBD for the entire 27 weeks, allowing for evaluation of longer-term drug effects.

The primary outcome is change in irritability subscale score of the Aberrant Behavior Checklist – 2nd edition (ABC-2) after 12 weeks of treatment.[41] The ABC-2 irritability subscale was selected as it was used in the FDA approval of aripiprazole and risperidone in the treatment of irritability and aggression in ASD.[4,5] Secondary aims compare CBD to placebo in the domains of anxiety, social skills, repetitive behaviors, executive functioning, sleep, parenting stress, quality of life, and overall autism symptoms. Each of these domains is assessed using validated scales for evaluation of children and adolescents with ASD (Table 1). The third aim evaluates for safety and adverse effects in youth with ASD treated with CBD compared to placebo.

**Table 1.**
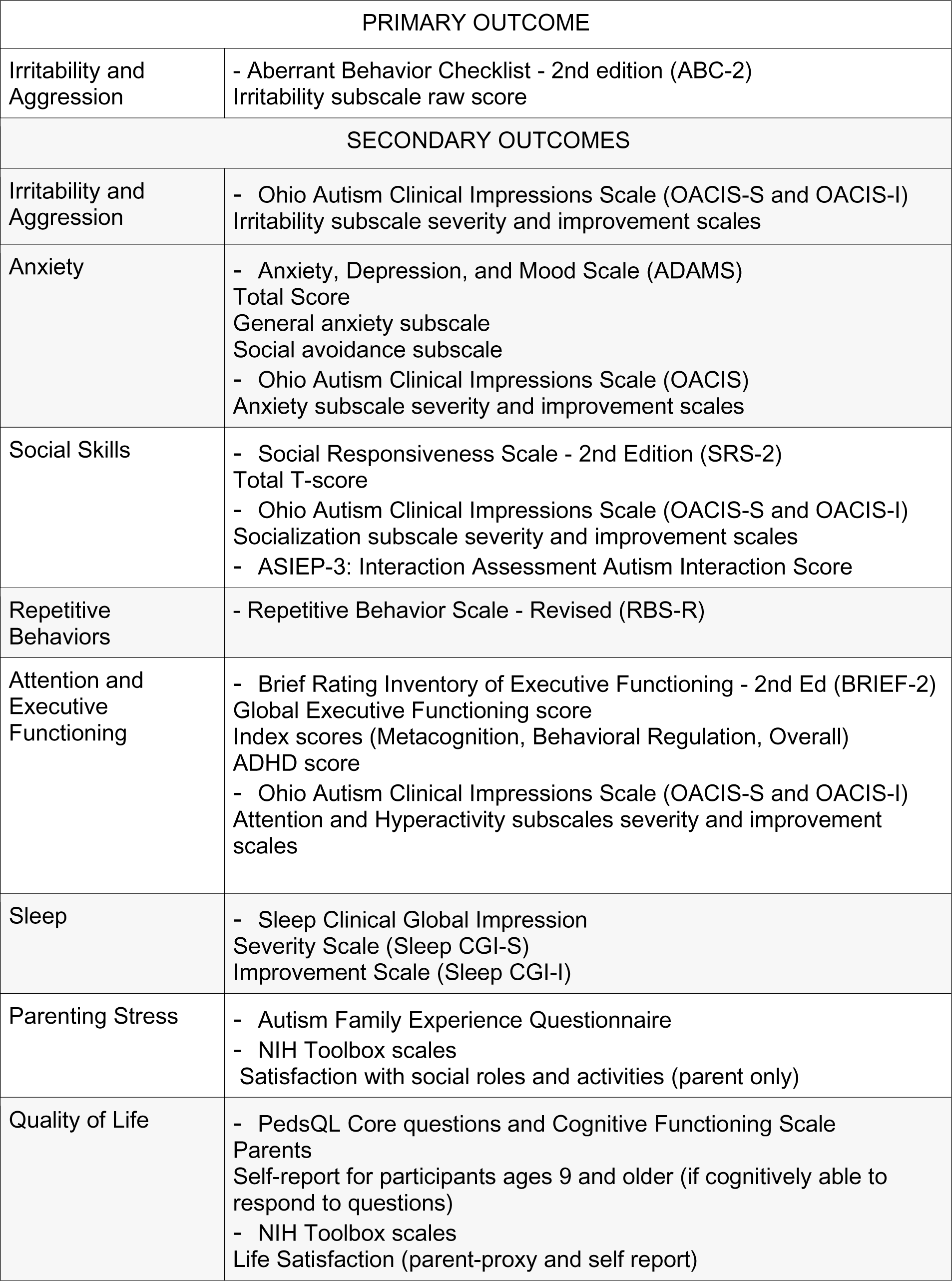
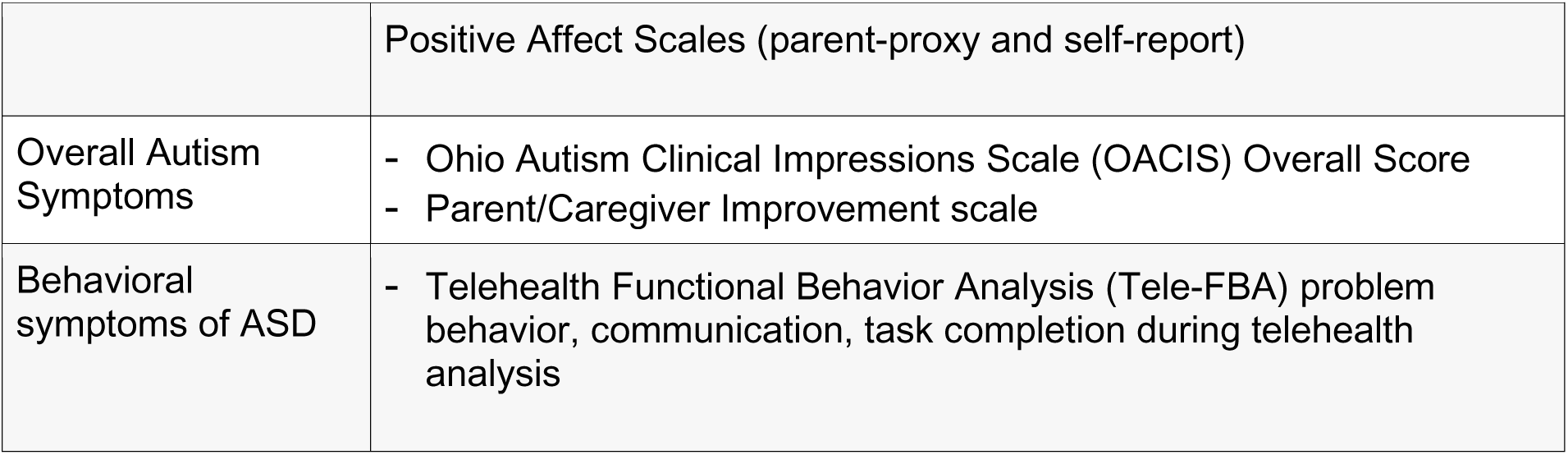
Summary of outcome measures.

Beyond these three aims, a small sample of participants receives CBD for the entire 27 weeks to provide pilot information on its continuing efficacy and safety (i.e. sustainability of CBD effects).

### Study Participants

Participants are recruited from a variety of locations, including Children’s Hospital Colorado clinics with high volumes of individuals with ASD and organizations in Colorado that support families with ASD. The study is also listed on clinicaltrials.gov (NCT04520685) and approved by the Colorado Multiple Institution Review Board (COMIRB 19-2168).

Inclusion criteria include healthy individuals aged 5-17 years with a previous diagnosis of ASD and a baseline ABC-2 irritability subscale score of at least 12. Participants can take up to two psychopharmacological medications and participate in behavioral therapies if there have been no changes for 4 weeks and no changes are planned during study. Certain seizure medications which impact the metabolism of CBD are exclusionary, as well as severe, unstable or progressive psychiatric symptoms. Full inclusion and exclusion criteria are listed in Table 2.

**Table 2.** Eligibility Criteria.

| Inclusion Criteria | Exclusion Criteria |
| --- | --- |
| 1) Children and adolescents aged 5-17 years | 1) Pregnant, nursing, or planning a pregnancy. Unwilling or unable to use standard acceptable methods of contraception for duration and one month after the last dose of study medication. |
| 2) In good health based upon results of medical history, physical examination, 12-lead ECG, and clinical laboratory tests. | 2) History of significant allergic condition, significant drug-related hypersensitivity, or allergic reaction to any compound or chemical class related to Epidiolex ®. |
| 3) Previous documented diagnosis of ASD | 3) Exposure to any investigational drug or device < 30 days prior to screening or at any time during the study. |
| 4) ABC-2 Irritability subscale $\geq 12$ at screening. | 4) Use of any THC or CBD-containing product within 4 weeks of Screening Visit, planned use during the study, or positive THC urine test at screening. |
| 5) OACIS-S Autism Global Rating Scale score $\geq 3$ at screening. | 5) Using the following medications: clobazam (Onfi, Frisium), felbamate (Felbatol), vigabatrin (Sabril), or everolimus (Afinitor). |
| 6) Stable regimen of $\leq 2$ psychiatric medications for $\geq 4$ weeks preceding study screening. Must maintain regimen throughout study. As needed medications for procedures not counted. | 6) Plan to initiate or change pharmacologic or non-pharmacologic interventions during the study. |
| 7) For patients with history of a seizure disorder: a) must currently be on a stable regimen of 1-2 anti-epileptic drugs, or b) must be seizure-free for 1 year if not currently receiving anti-epileptic drugs. | 7) ALT, AST, total bilirubin, or creatinine levels $\geq 2$ times the upper limit of normal, alkaline phosphatase levels $\geq 3$ times the upper limit of normal, or hematocrit $< 1.2$ times lower limit of normal. |
| 8) For patients with history of a seizure disorder: must be on a stable AED regimen for 3 months prior to and throughout study. | 8) Severe or unstable symptoms of ASD |
| 9) Non-pharmacological educational, behavioral, and/or dietary interventions or therapies must be stable for $\geq 2$ months. | 9) Acute or progressive neurological disease or psychiatric disorder |
| 10) Body mass index between 12-32 kg/m<sup>2</sup>. | 10) Suspected or confirmed cardiovascular disease. |
| 11) Females of childbearing potential must have negative serum pregnancy test at screening and all designated visits. | 11) History of treatment for, or evidence of, drug or alcohol abuse within the past year. |
| 12) Agree to abide by all study restrictions and procedures. |  |
| 13) Able to read and respond to questions and questionnaires in English. |  |
| 14) Adequately informed of the nature and risks of the study. |  |
| 15) Written consent to assist in study drug administration. |  |
| 16) Reliable and willing and able to comply with all protocol requirements and procedures. |  |

### Randomization and Blinding

After screening assessments eligible participants are randomized. A permuted block randomization scheme was generated to randomly assign participants to one of the 3 study arms: CBD/placebo (Arm A), placebo/CBD (Arm B), or CBD/CBD (Arm C). Because the CBD/CBD group was designed to have 1/3 as many participants as the other two groups, 2/3 of the assignments to the CBD/CBD group were sequentially deleted from the randomization scheme, leaving only 12 participants in that group. The pharmacist then translated the result into a final randomization list so that all other investigators, participants, their parents/caregivers, study coordinators, and examiners are blind to randomized group assignment. During the study, the blind is broken only in emergencies when knowledge of the patient’s treatment group is necessary for further management, e.g. severe adverse side effects. See Figure 1 for study design.

**Figure 1:**
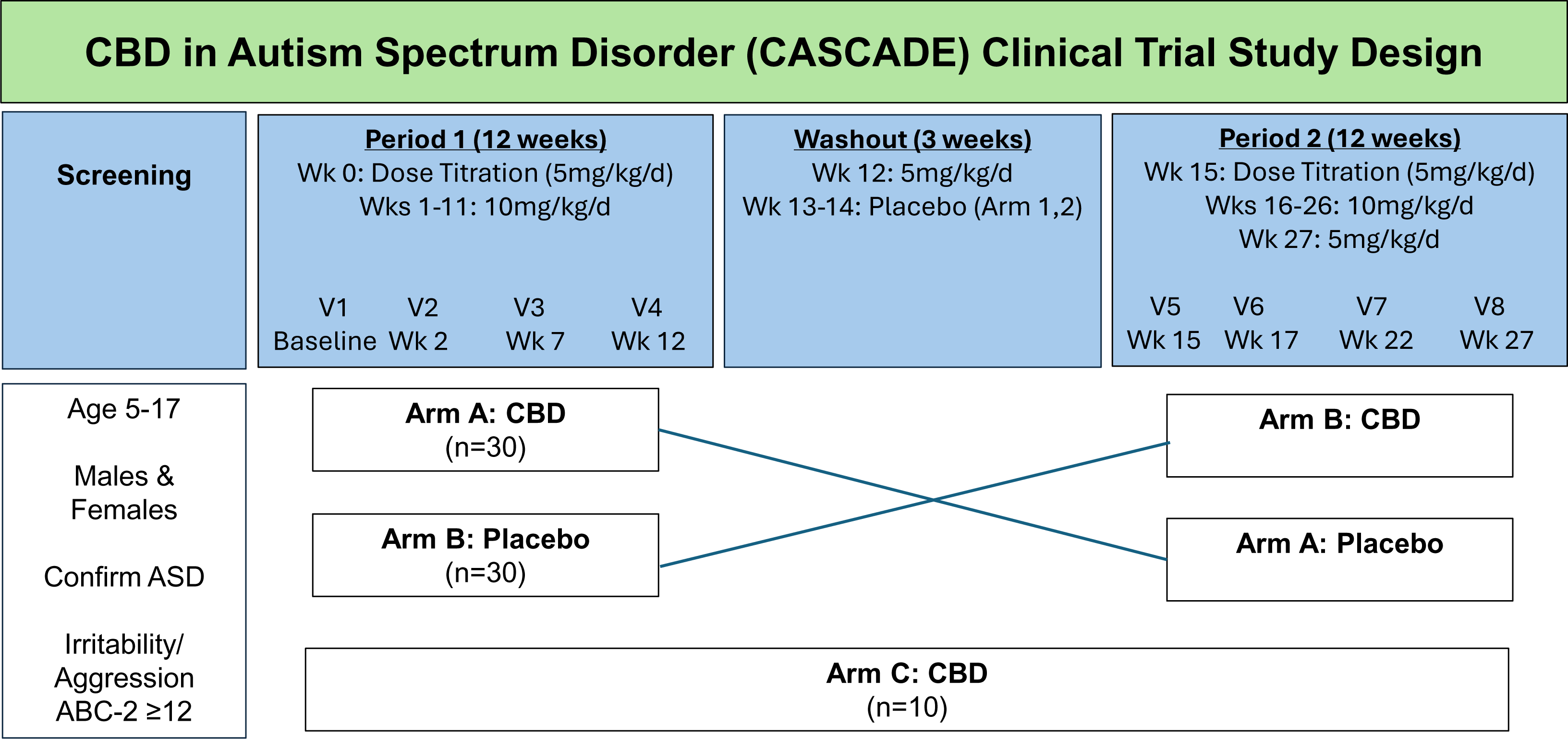
CASCADE Study Design.

### Intervention

The investigational product is cannabidiol 100mg/mL liquid (Epidiolex ®; Jazz Pharmaceuticals). Placebo contains identical packaging and ingredients minus the CBD. Bottles labelled Bottle A and Bottle B are dispensed at the beginning of Period 1 and Period 2, and dosing instructions are provided. Caregivers of participants receive training on administration of the study drug and are assessed in their ability to draw up the prescribed volume, as well as proper storage and handling. Daily dosing diaries are completed by caregivers.

During the periods of CBD treatment, the dose is started at 5mg/kg/day divided twice per day, increased to 10 mg/kg/day divided twice per day after 1 week, and continued for 11 weeks for Arms A and B and for 26 weeks for Arm C (see Figure 1). Following CBD treatment, the dose decreases to 5mg/kg/day for 1 week and is then discontinued. Blinding is maintained by administering the same volume of medication from 2 different bottles labelled Bottle 1 and Bottle 2, with specific instructions provided by the research pharmacy for dosing from each bottle. For all 3 arms, a dose decrease to 5mg/kg/day is allowed if adverse effects occur that are possibly related to study drug. If adverse effects do not resolve with the decreased dose, the participant is discontinued from the study.

### Study Visits and Assessments

At the screening visit, documentation of previous ASD diagnosis is reviewed and confirmed. A confirmatory evaluation for autism diagnosis is conducted using an ASD DSM-5 diagnostic criteria checklist[1] and administration of the Autism Diagnostic Observation Schedules (ADOS-2).[42] If the participant does not meet criteria for ASD based on DSM-5 criteria, they are not eligible for participation. Additional baseline assessments include the Stanford Binet Intelligence Scale (SB-5 ABIQ)[43] and Vineland Adaptive Behavior Scales – 3^rd^ Edition.[44] Cognitive and adaptive scores are utilized to characterize the study population, not as study endpoints.

Medical and developmental history are collected at screening including demographics, medical and/or genetic diagnoses, current and previous neurodevelopmental diagnoses and common comorbidities (such as attentional disorders, intellectual disability, or anxiety disorders), current and previous medication use, educational setting and current therapies. Subsequent visits inquire about any changes to these variables or other important life changes affecting the participant.

The study schedule includes in-person visits at screening, baseline (week 0), week 2, week 7, week 12 (end of Period 1), week 15 (start of Period 2), week 17, week 22, and week 27. In most cases, screening and baseline (week 0) visits are combined into a single visit unless the participant prefers separate visits. Phone visits occur at week 1, week 4, week 16, week 19, and week 30. Outcome measures (listed below and in Table 2) are collected at baseline, week 7, and week 12 for both Period 1 and Period 2. All in-person visits include vital signs, physical examination, adverse event monitoring, review of the medication diary, measurement of study product for monitoring compliance, blood draw for safety laboratory analyses and compliance, and suicidality screening if developmentally appropriate. Phone visits are conducted to monitor tolerability, adverse events, screen for suicidality, and to answer any additional questions about the study protocol. Early termination visits are scheduled prior to discontinuation of study drug if a participant is going to withdraw from the study, unless the study drug needs to be stopped for safety concerns prior to the early termination visit. A post-study computer survey collected 4 weeks after study completion collects feedback about experiences of study participation, asks parents their perception of the study arm assignment, and collects further input or plans related to future CBD use for their child.

Social stories and visual schedules including photos of study staff, locations in the research clinic, and procedures are provided to caretakers prior to visits for participant preparation. Reinforcers are utilized to improve motivation and participation. Support and discussion about behavioral or psychosocial concerns with study clinicians (Pediatrics, Psychiatry, Psychology) is available to parents if needed to determine if study participation is appropriate compared to more intensive clinical care.

### Baseline Assessments

#### ASD DSM-5 Checklist

The ASD DSM-5 Checklist contains the diagnostic criteria for ASD including the domains of social communication and restricted, repetitive behaviors and confirms eligibility for the study based on diagnosis.[1]

#### Autism Diagnostic Observation Schedule (ADOS-2)

The ADOS-2 is a semi-structured play-based assessment aimed at eliciting core features of autism spectrum disorder, including social and language responses expected at particular developmental levels. It is considered part of the gold-standard evaluation of autism spectrum disorder. [42,43]

#### Stanford-Binet Intelligence Scales – Abbreviated IQ (SB-5 ABIQ)

The SB-5 is a commonly accepted standardized measure of cognitive skills, with a set of subtests that have been identified as a reliable estimate of IQ which make up the ABIQ (Abbreviated IQ) score. Children with ASD may have a range of cognitive abilities, from low functioning in the intellectual disability range to giftedness. This profile within the sample is important for data analysis and evaluating treatment response.[44,45]

#### Vineland Adaptive Behavior Scales (Vineland-3)

The Vineland-3 Adaptive Behavior Scales aids in diagnosing and classifying disability in different domains of adaptive functioning including communication, daily living skills, socialization, and motor skills.[46–48]

### Outcome Measures

#### Aberrant Behavior Checklist – 2 (ABC-2)

The ABC-2 is a symptom checklist assessing problem behaviors in individuals with intellectual disability across settings. The five subscales are: irritability/aggression, hyperactivity, social avoidance, stereotypy, and inappropriate speech.[41,49–51] Change in the irritability/aggression subscale score is the primary outcome measure of the study.

#### Anxiety, Depression and Mood Scale (ADAMS)

The ADAMS is parent-report scale designed for individuals with autism and other disabilities to measure domains of anxiety, depression, and mood. It has demonstrated good internal consistency and test-retest reliability.[52,53]

#### OACIS-S (Ohio Autism Clinical Impressions Scale – Severity) and OACIS-I (Ohio Autism Clinical Impressions Scale – Improvement)

The OACIS-S and OACIS-I assess severity and improvements, in domains of social interaction, aberrant behavior (including irritability and aggressive behavior), repetitive or ritualistic behavior, verbal communication, nonverbal communication skills, hyperactivity and inattention, anxiety and fearfulness, sensory sensitivities, restricted and narrow interests, and a global rating of autism severity.[54,55]

#### Social Responsiveness Scale – 2^nd^ Edition (SRS-2)

The SRS-2 is a standardized parent-report questionnaire that provides T-scores in domains of communication, motivation, cognition, and restrictive/repetitive behaviors.[56,57]

#### Autism Screening Instrument for Educational Planning – 3 (ASIEP-3)

The ASIEP is a video-taped play-based assessment that evaluates social interactions and social responses.[58,59]

#### Repetitive Behavior Scale-Revised (RBS-R)

The RBS-R is a caregiver questionnaire assessing repetitive behaviors with 6 subscales: stereotyped, ritualistic, self-injurious, compulsive, restricted, and sameness.[60,61]

#### Pediatric Sleep Clinical Global Impressions Scales

The Pediatric Sleep CGI scales measure pediatric insomnia in ASD and have been validated against the gold-standard sleep questionnaire and actigraphy. It includes a structured sleep history form, with questions about falling and remaining asleep independently; bedtime resistance; sleep onset delay; night awakening; caregiver satisfaction with the child’s sleep patterns; and family functioning, as impacted by the child’s sleep patterns.[62]

#### Behavioral Rating Inventory of Executive Functioning (BRIEF-2)

The BRIEF-2 is a standardized rating scale that measures domains of executive functioning, including behavior regulation (inhibition, self-monitoring), emotional regulation (shifting, emotional control), and cognitive regulation (initiation, working memory, plan/organize, task monitoring, and organization of materials).[63,64]

#### Autism Family Experience Questionnaire (AFEQ)

The AFEQ measures family quality of life with domains specific to raising a child with ASD, the child’s development and wellbeing, and family functioning. The AFEQ is an ecologically validated instrument, shown to have excellent internal consistency and sensitivity to change.[65]

#### NIH Toolbox Measures

National Institutes of Health Patient Reported Outcome Measures Information System (NIH PROMIS) is used to quantify psychological wellbeing and quality of life. Assessments in the study include life satisfaction, positive affect, and satisfaction with participation in social roles. Psychometric results of NIH PROMIS indicate excellent reliability and convergent validity.[66–68]

#### Pediatric Quality of Life Inventory (PedsQL)

The PedsQL is a caregiver questionnaire to evaluate overall quality of life in different domains.[69,70]

#### Telehealth Functional Behavior Analysis (Tele-FBA)

The Tele-FBA protocol is an optional assessment in which a telehealth functional analysis is performed to evaluate behavior in natural settings. In the Tele-FBA session, common reinforcers for problem behavior (access to attention/tangibles; escape from demands) are evaluated in a series of test and control conditions, each lasting 5 min. Sessions are conducted within 3 days of the study visits at baseline, week 10-11, and at week 25-26.

### Safety Monitoring

Safety monitoring includes vital signs (heart rate, blood pressure, height, and weight), physical examination, laboratory tests, adverse medical or behavioral symptoms, and screening for suicidality (as evaluated via the Columbia Suicide Severity Rating Scale, C-SSRS)[71,72].

Electrocardiogram is obtained at baseline. Laboratory tests collected at each in-person visit include comprehensive metabolic profile (electrolytes; AST and ALT; total bilirubin; BUN and creatinine), complete blood count (white blood cell count with differential; hemoglobin and hematocrit; platelet count), and hemoglobin A1c. Blood samples collected also include analysis of CBD levels, CBD metabolites, THC, and other cannabinoid levels, as well as optional biobanking for future genetic and metabolic studies. Urine pregnancy test is conducted at each visit for pubertal females. Adverse medical and behavioral symptoms are recorded at each visit and phone call in regards to incidence, nature, duration, intensity, potential relationship with study drug, additional treatments, and any adjustments to study drug dosing.

### Statistical and Power Analysis

Power Analyses: Using SAS Proc Power and reported estimates on the ABC-2 irritability subscale score [73], we have specified a significance level of alpha= 0.05, two-tailed, and allowed for increased variability to determine the minimum difference between groups detectable with 70 subjects, where 40 receive CBD and 30 cross over to placebo. Note that for the 10 patients who do not cross over to placebo (those providing sustainability information), their data during the first CBD phase can still be included in the mixed model analyses. Even though data for the placebo phase is missing for these participants, the models allow for missing data, assuming data are missing at random. As a simplistic and likely conservative estimate of power based on a paired t-test that does not account for covariates and assumes no period effect, this design provides 80% power to detect a 5.75 point or greater difference on the ABC-2 irritability subscale between CBD and placebo groups and 80% power to detect a treatment response rate (OACIS Total Autism Score-I of 1 or 2) in the CBD group of 52% or greater, assuming a 20% response rate in the placebo group.

Analyses: Primary and secondary outcomes for Aims 1 and 2 are analyzed for the intent to treat sample (i.e. all randomized subjects) with linear and generalized linear mixed models that can include participants with missing data; the potential period effects (i.e. order effects) are evaluated and included if significant. Models specify condition (CBD, placebo), time (0, 7, 12 weeks) and the condition by time interaction as fixed effects and subject as a random effect, where subject is assumed independently normally distributed with mean 0 and variance independent of the random errors and assuming an unstructured covariance. These are mixed model analyses of covariance (ANCOVAs) where means are estimated at each time point and are on the link function scale when general linear mixed models are utilized. A significance level of alpha=0.05 is used to evaluate a priori planned comparisons. Closed testing procedures with hierarchical evaluation provide some protection against multiple comparisons. Tests for group differences at each time are conducted only for those outcomes with significant group by time interactions. Efficacy is determined by significantly greater improvement in irritability score in the CBD group vs placebo at week 12. Secondary models will evaluate variables such as sex, age, IQ and autism severity as potential covariates and report any differences in results compared to the primary models. To address attrition, sensitivity analyses are conducted as appropriate and/or results are compared with and without incorporating multiple imputed values from a generalized linear mixed model approach for missing data.

Preliminary evidence of sustainability of outcomes is evaluated via visual and descriptive comparisons of mean scores throughout both 12-week periods (including washout) for the 10 patients who continue CBD for both study periods. For each patient, we evaluate the reduction in ABC-2 irritability subscale score (and secondary outcomes) after the first 12-week period and consider maintenance of at least 70% of that reduction during the second period on CBD as preliminarily evidence of sustainability.

Rates of adverse events, treatment response, and the pattern of missing data are evaluated with logistic and/or binomial regressions comparing the CBD and placebo phases that account for repeated measures on subjects.

### Ethics

The study is conducted according to the Declaration of Helsinki, Protection of Human Volunteers (21 CFR 50), Institutional Review Boards (21 CFR 56), and Obligations of Clinical Investigators (21 CFR 312). Review and approval was provided by Colorado Multiple Institution Review Board (COMIRB; Protocol 19-2168). The study population is vulnerable because it includes children, many of whom have cognitive impairment. Because of the vulnerable nature of the participants, all informed consent and assent meetings are facilitated by one of the study clinicians specialized in ASD who can answer specific questions related to the study medication and other treatment options.

### Limitations

Limitations of the study include limited treatment duration for evaluation of long-term effects and sample size with insufficient power to analyze drug effect in regards to sex and race/ethnicity differences, varying levels of ASD symptoms and intellectual function. Further, allowing participants with concomitant psychiatric medications decreases the ability to study the effect of CBD alone on baseline symptoms, leading to potential underestimation of efficacy as the other medications are already addressing some symptoms. While drug naïve participants are included, duration of time for recruitment of an entirely drug naïve cohort is limiting. Further, life changes and other unanticipated stressors (big or small) can lead to behavioral changes for some individuals with ASD, and while randomized and enrolled at different timepoints throughout the school year, the impact of life events on behavioral outcomes may differ between groups. There is also risk for placebo effect as this is a vulnerable and often challenging population with caregivers heavily invested in potential treatments leading to behavioral improvements.

## CONCLUSION

There is a great need for safe medications to address irritability, aggression and other challenging behavioral symptoms associated with autism spectrum disorder in children. FDA approval for aripiprazole and risperidone occurred almost 15 years ago (2009 and 2006, respectively) and there continues to be a dearth of available, effective, and well tolerated medication options to treat this population. While still limited, research to date on use of cannabinoid products including CBD in autism have demonstrated promise in improving negative behavioral symptoms, while also showing a relatively strong safety profile.[74,75] Thus, there is clear justification for additional placebo-controlled trials with larger sample sizes such as the CASCADE study, as well as additional trials with longer follow-up.

To our knowledge, there are currently 4 other active or recruiting placebo-controlled studies investigating the effects of CBD on children and adolescents with ASD which will also contribute to answering this question.[75] If positive, further trials comparing efficacy to current treatments as well as studies evaluating CBD in combination with evidence-based behavioral treatments are important. However, publication of negative study results is also important, as the ASD community in general is historically and currently plagued by trends of different anecdotal and non-evidence based treatments that can lead to unnecessary expense, wasted time, emotional disappointment, and risks to health and safety of autistic individuals and their families.

Determination of the safety and efficacy of CBD for treatment of irritability and aggression in individuals with ASD is vital, as many families are already using cannabidiol products to treat their loved ones.

## Data Availability

All data produced in the present study are available upon reasonable request to the authors following peer-reviewed publication

## ACKNOWLEDGEMENTS

We acknowledge past research coordinators and study team members participating in this study including Steffany Contreras, Nicole Semmler, Nana Welnick, Eva Adana, Sailor Brukardt, Amy Taylor, and Hannah Fleming. We also acknowledge Children’s Hospital Colorado CTRC Nursing Team.

## CRediT AUTHOR STATEMENT

**Elise M. Sannar:** Conceptualization, Methodology, Investigation, Writing – Reviewing and Editing, **Joan R. Winter:** Writing – Original Draft, **Ronda K. Franke**: Investigation, **Emily Werner:** Investigation, **Rebecca Rochowiak:** Data curation, Resources, **Patrick W. Romani:** Investigation**, Owen S. Miller:** Investigation, **Jacquelyn L. Bainbridge:** Conceptualization, Methodology, Supervision, **Talia Thompson:** Investigation, **Crystal Natvig:** Data Curation, Formal analysis, **Susan K. Mikulich** – Conceptualization, Methodology, Formal analysis, Writing – Reviewing and Editing, **Nicole R. Tartaglia** – Conceptualization, Methodology, Investigation, Writing – Reviewing and Editing, Supervision, Project administration, Funding acquisition.

